# Characterizing the impact of plasma protein levels on human brain structure and disorders leveraging integrative multi-omics analysis

**DOI:** 10.64898/2026.07.13.26358006

**Authors:** Cyrus Ayubcha, Evan Dennis, Upasana Bhattacharyya, Jibin John, Max Lam, Todd Lencz, Tian Ge, Chia-Yen Chen

**Author notes:** These authors jointly supervised this work: Chia-Yen Chen; Tain Ge.

## Abstract

With recent advances in high-throughput proteomic technologies, population-scale plasma proteomics datasets, often linked to extensive genetic and phenotypic information, have become increasingly accessible. Yet the relationships between circulating protein levels, brain imaging phenotypes, and risk for neurological and psychiatric disorders remain largely unexplored. Proteome-wide association studies offer a promising approach for elucidating biological mechanisms that connect genetic variation to complex brain-related traits and diseases. In this study, we integrated protein quantitative trait loci (pQTLs) from the two largest plasma proteomic resources (the UK Biobank Pharma Proteomics Project [UKB-PPP] and Ferkingstad *et al.* [deCODE]) with genome-wide association studies of brain imaging-derived phenotypes in UK Biobank using Mendelian randomization and colocalization analyses. We identified 120 cis and 20 trans associations between plasma proteins and imaging phenotypes and validated these findings using brain tissue-derived proteomic and transcriptomic datasets. Multivariable Mendelian randomization revealed eleven plasma proteins (coding genes *APOE, ARL3, MICB, NSF, RHOC, RSPO3, ENPP2, BTN2A1, EIF2AK3, MRVI1,* and *OPLAH*) with significant direct effects on the risk of Alzheimer’s disease, Parkinson’s disease, multiple sclerosis, bipolar disorder, and schizophrenia. Single-cell expression and pathway enrichment analyses further revealed cell-type-specific effects and distinct biological processes underlying these protein-disease associations. Together, these findings demonstrate robust links between plasma protein variation and brain structure, delineate protein-disease pathways, and highlight the cellular and molecular mechanisms that contribute to neurobiological diversity and pathology.

## Introduction

Large-scale genetic association studies in human populations have long provided insight into the genetic architecture of disease, informing target discovery, pharmacologic repurposing, and patient stratification. However, interpreting how genetic variation gives rise to complex disease phenotypes remains challenging, in part because most findings reflect statistical associations embedded within a highly intricate genomic landscape.^1^

Consequently, recent research has shifted toward characterizing downstream molecular mechanisms, particularly how genetic variants influence RNA and protein abundance through expression quantitative trait loci (eQTLs) and protein quantitative trait loci (pQTLs).^2,3^ Although early QTL studies focused primarily on cis-QTLs (typically variants located within 500-1,000 kb of the gene), trans-QTLs (variants located beyond this proximal window) have become increasingly recognized for their biological relevance, especially in pQTL studies.^2,3^ Trans- pQTLs often act through intermediate genes, signaling cascades, or transcriptional regulators to modulate distal protein expression.^3^ Together, large-scale eQTL and pQTL resources provide systems-level insight into the mechanisms linking genetic variation to phenotypic diversity.^4^

Two of the largest plasma pQTL datasets available were generated by Ferkingstad *et al.* (referred to as the deCODE study hereafter) and the UK Biobank Pharma Proteomics Project (UKB-PPP). The deCODE study quantified 4,719 plasma proteins in 35,559 Icelanders using the SomaScan multiplex aptamer assay, identifying 18,084 significant pQTLs, including 1,881 cis and 16,203 trans associations, across 4,631 proteins.^5^ The UKB-PPP consortium profiled 2,942 plasma protein analytes in 34,557 randomly selected UKB participants using the Olink Explore 3072 platform, mapping 14,287 significant pQTLs from 3,760 independent genetic regions, including 1,955 cis and 12,332 trans associations.^6^ A recent comparative analysis highlighted complementary strengths across the two platforms: SomaScan showed higher assay precision, whereas Olink detected a larger number of cis-pQTLs; both revealed demographic and disease-specific variability in pQTL detection, as well as substantial pleiotropy, with some genetic variants influencing multiple proteins.^7^

pQTLs have increasingly been used as genetic instruments in Mendelian Randomization (MR)^8^, which leverages the random allocation of alleles at conception to estimate causal effects under three key assumptions: independence (instruments are unconfounded), relevance (instruments are strongly associated with the exposure), and exclusion restriction (instruments influence the outcome only through the exposure).^9^ Guided by these principles, several studies have applied MR to pQTLs derived from brain tissue to investigate potential causal relationships between protein abundance and neurological disorders such as stroke, Alzheimer’s disease (AD), and Parkinson’s disease (PD).^10,11^

Most MR studies to date have focused on disease endpoints. In contrast, neuroimaging phenotypes provide quantitative biomarkers that capture fundamental pathophysiological processes underlying neurological and psychiatric disorders.^12,13^ The UKB hosts one of the largest single-cohort neuroimaging datasets, comprising nearly 4,000 brain imaging-derived phenotypes (IDPs) from approximately 40,000 participants with accompanying genome-wide genotypes.^14^ Recent UKB GWAS of IDPs have identified numerous genetic loci associated with gray- and white-matter structure, regional brain volumes, and other neuroanatomical traits.^14,15^

In this study, we integrate pQTLs from large-scale plasma proteomic datasets with UKB neuroimaging GWAS to investigate how genetically predicted plasma protein abundance relates to brain structure and organization. Using MR and complementary multi-omics analyses, we (i) identify gene-protein-imaging pathways, (ii) validate associations using brain-derived transcriptomic and proteomic datasets, (iii) perform mediation analyses across multiple neurological and psychiatric disorders, and (iv) characterize cell-type-specific and pathway-level mechanisms. Collectively, our results provide a systematic framework for linking plasma protein variation to human neuroimaging phenotypes and illuminate potential biological pathways underlying neurological and psychiatric disease.

## Results

### Plasma cis-pQTL and trans-pQTL Instruments for Mendelian Randomization

The analytic workflow is summarized in **Supplementary Fig. 1**. We first curated and quality-controlled pQTL summary statistics from both UKB-PPP (Sun *et al.* 2023)^6^, generated using the Olink Explore assay, and from the deCODE study (Ferkingstad *et al.* 2021)^5^, generated using the SomaScan platform, to derive genetic instruments for MR. From UKB-PPP, we obtained 17,138 genome-wide significant cis-pQTLs associated with 2,020 unique proteins measured by Olink. From the deCODE dataset, we identified 23,348 genome-wide significant cis-pQTLs associated with 1,622 unique proteins measured by SomaScan. For trans effects, we curated 31,622 trans-pQTL instruments (excluding the extended MHC regions) corresponding to 2,529 unique proteins in UKB-PPP, and 72,360 trans-pQTLs associated with 4,192 unique proteins in the deCODE dataset. Because many proteins were represented across multiple datasets and QTL sources, the combined instrument set encompassed 5,310 unique plasma proteins aggregated across the four pQTL collections (**Supplementary Fig. 2; Supplementary Table 1**).

### Investigating the Effects of Plasma Protein Levels on Brain Imaging Phenotypes

We first examined the effects of plasma cis- and trans-pQTLs on 556 selected brain imaging-derived phenotypes (IDPs; **Supplementary Table 2**), including subcortical volumes, cortical thickness and surface area measures, and features derived from diffusion tensor imaging (DTI), using Mendelian randomization (MR). Using cis-pQTL instruments from the UKB-PPP and deCODE datasets, we identified 600 unique significant protein-IDP associations, where specific protein-IDP pairs could be implicated across multiple MR methods. Of these, 312 were detected exclusively using UKB-PPP instruments, 242 were detected only using deCODE instruments, and 46 associations were replicated across both databases (Bonferroni-corrected thresholds: UKB-PPP P < 4.48×10^−8^; deCODE cis-pQTL *P* < 5.41×10^−8^; **Figure 1; Supplementary Table 3**). Trans-pQTL MR analysis identified an additional 20 significant associations using the weighted median (WM) method (Bonferroni-corrected thresholds: UKB-PPP 3.54×10^−8^; deCODE 1.91×10^−8^; **Supplementary Table 4**). **Table 1** summarizes all significant cis-pQTL and trans-pQTL MR findings.

**Figure 1.**
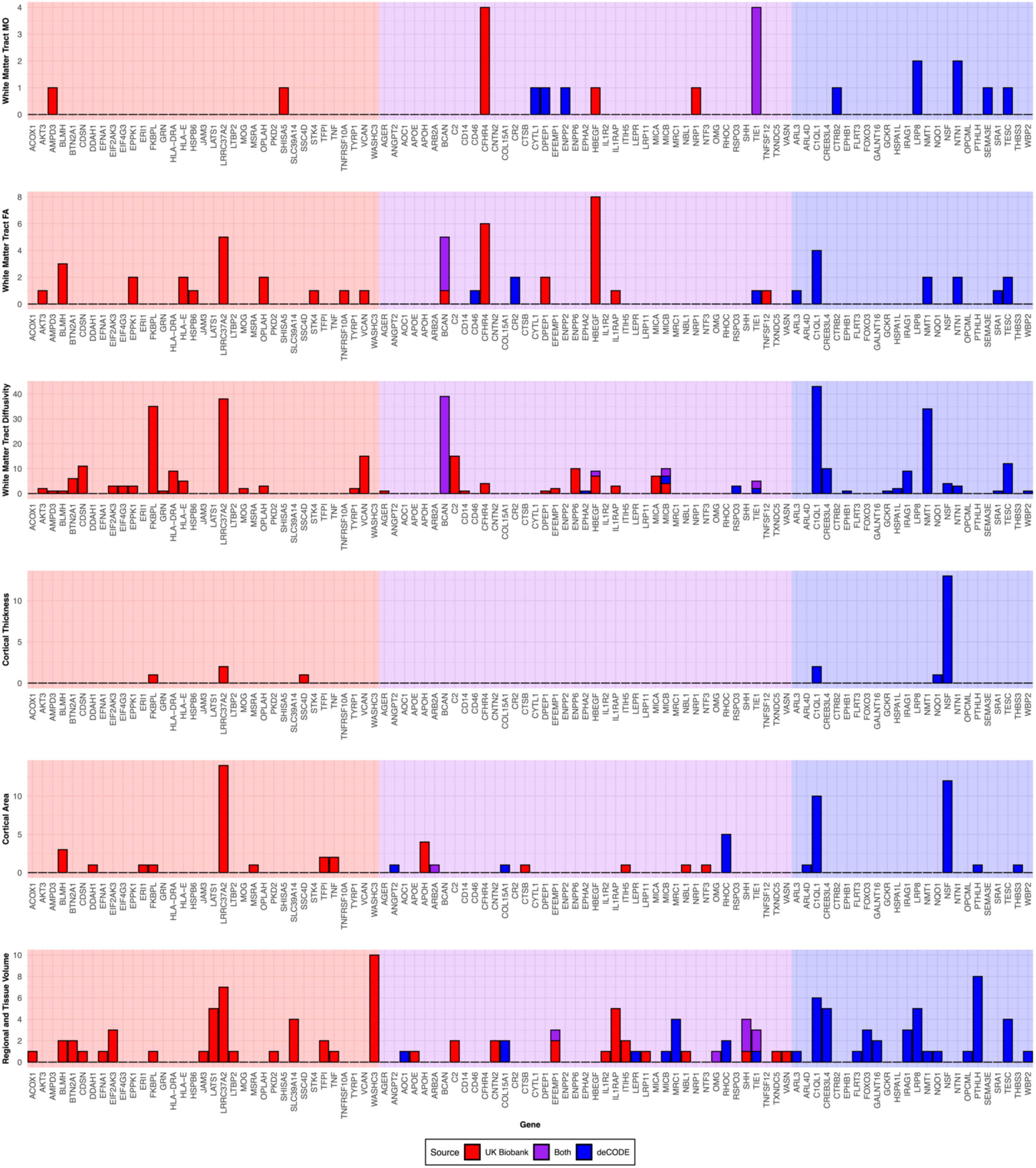
Summary of significant cis-pQTL effects on IDPs identified through Mendelian randomization. The figure displays the number of significant plasma protein-IDP associations identified in the primary Mendelian randomization analysis, stratified across six neuroimaging modalities and measurement types.

**Table 1.** Summary of Mendelian randomization (MR) results for imaging-derived phenotypes using cis- and trans-pQTLs from the UK Biobank Pharma Proteomics Project (UKB-PPP) and the deCODE study.

|  | pQTL:<br>MR method | Total number<br>of tests | Number of<br>proteins<br>analyzed | Number of<br>significant<br>MR findings | Number of unique<br>proteins in<br>significant findings | Number of unique<br>IDPs in significant<br>findings |
| --- | --- | --- | --- | --- | --- | --- |
| UKB-PPP | Cis: Wald ratio | 1,129,236 | 2,031 | 309 | 57 | 164 |
|  | Cis: WM | 669,760 | 969 | 18 | 6 | 18 |
|  | Cis: IVW | 868,894 | 1,167 | 70 | 11 | 61 |
|  | Trans: WM | 1,243,242 | 2,079 | 10 | 8 | 8 |
| deCODE | Cis: Wald ratio | 929,076 | 1,167 | 286 | 42 | 195 |
|  | Cis: WM | 579,462 | 1,120 | 9 | 3 | 10 |
|  | Cis: IVW | 697,866 | 1,453 | 11 | 7 | 11 |
|  | Trans: WM | 1,356,862 | 2,269 | 10 | 6 | 10 |

IDP: imaging derived phenotype; WM: weighted median MR estimate; IVW: inverse-variance weighted MR estimate.

To validate the cis-pQTL MR findings, we applied both conventional Bayesian colocalization (coloc-abf)^16^ and Sum of Single Effects fine-mapping with colocalization (SuSiE-coloc)^17^ to assess whether cis-pQTL instruments colocalize with IDP-associated genetic signals. SuSiE-coloc produced credible sets for 149 cis-pQTL-IDP pairs in UKB-PPP and 93 in deCODE. Across all 600 significant cis-pQTL-IDP associations, 120 pairs spanning 28 unique proteins exceeded the posterior probability threshold for colocalization (H4 > 0.7), with some pairs supported by both colocalization methods (**Figure 2a; Supplementary Table 5**). Within UKB-PPP, 88 of 358 significant MR pairs (312 UKB-PPP only and 46 replicated; 88/358 = 25%) colocalized. Within deCODE, 59 of 288 significant MR pairs (242 deCODE only and 46 replicated; 59/288 = 20%) colocalized. Several proteins colocalized with multiple distinct IDPs, suggesting broader neuroanatomical influence. For example, *PTHLH* colocalized with two IDPs (**Figure 2b; Supplementary Table 5**).

**Figure 2:**
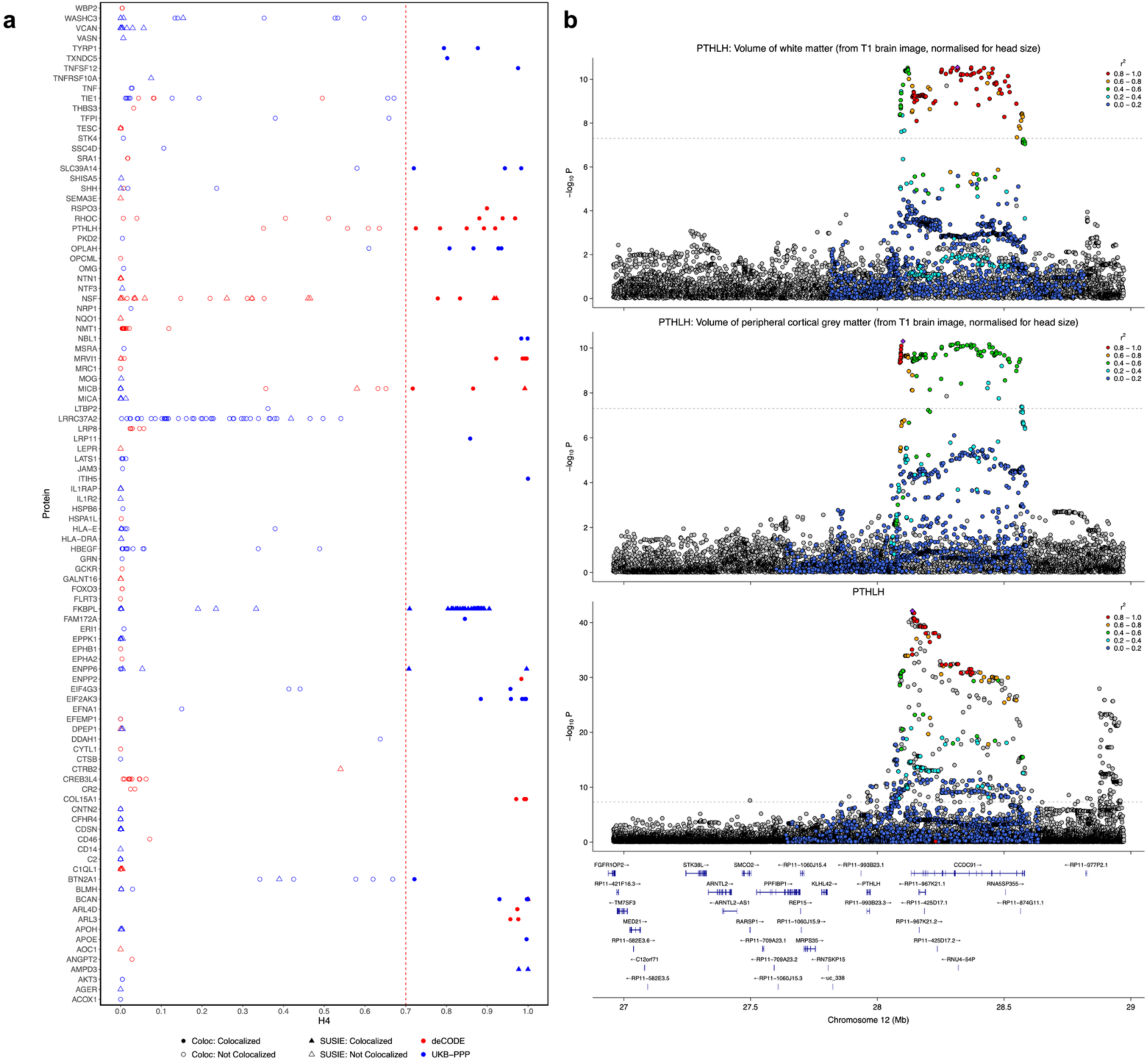
Colocalization analysis validating MR-identified plasma protein-IDP associations. (A) Summary of cis-pQTL-IDP pairs with strong evidence of colocalization, defined as the posterior probability of a shared causal variant (H4 > 0.7). “coloc” denotes results from the single causal variant model (coloc.abf), while “SuSiE” denotes the fine-mapping based approach (SuSiE-coloc). For pairs analyzed by both methods, the higher H4 value is shown. (B) Regional LocusZoom plots illustrating colocalized associations between *PTHLH* pQTLs and corresponding IDP GWAS signals (H4 > 0.7).

### Validating Plasma Proteomics Findings for Brain Imaging Phenotypes using Brain Tissue-Specific Data

To assess whether plasma protein associations with brain IDPs reflect underlying biology in neural tissues, we sought to replicate our plasma pQTL-based findings using brain tissue-derived transcriptomic and proteomic resources. We first performed MR analyses using pQTLs from the Religious Orders Study/Memory and Aging Project (ROSMAP) dataset (N = 706)^18^ for 41 proteins that showed either significant cis-pQTL colocalization or significant trans-pQTL weighted median MR effects in the plasma-based analyses. Of these 41 proteins, 20 were available in ROSMAP (**Supplementary Table 6**). However, SNP instruments for SEMA4C were missing from the IDP GWAS, leaving 19 proteins for evaluation. Using single-SNP MR (Wald ratio) with ROSMAP brain pQTL instruments, we assessed 65 unique protein-IDP pairs. Nearly 75% (49/65) replicated at Bonferroni significance (*P* < 0.05/65 = 7.69×10^−4^; **Supplementary Table 7**), supporting the robustness of the plasma-derived associations in brain tissue.

As an additional layer of validation, we conducted MR analyses using brain eQTLs from the MetaBrain dataset (N = 6,526)^2^. Among the 41 proteins implicated in the plasma analyses, 28 corresponding genes were represented in MetaBrain (**Supplementary Table 8**); however, only 17 could be tested due to missing SNP instruments in the IDP GWAS. Using MetaBrain brain eQTLs instruments, we evaluated 49 unique protein-IDP pairs and identified 22 significant associations (Bonferroni-corrected threshold: 0.05/49 = 1.02×10^−3^; **Supplementary Table 9**). The partial replication observed likely reflects biological and regulatory differences between transcriptomic and proteomic layers.

### Estimating Mediated Effects of Plasma Protein Levels on Neurological and Psychiatric Disorders through Brain Imaging Phenotypes

To investigate how plasma protein levels influence neurological and psychiatric disorders, particularly through their effects on brain structure, we applied multivariable Mendelian randomization (MVMR) for causal mediation analysis. This framework enabled estimation of the total, direct, and indirect (IDP-mediated) effects of plasma proteins on disease risk. Consistent with earlier validation analyses, we restricted the evaluation to plasma proteins that exhibited either significant cis-pQTL colocalization or significant trans-pQTL weighted median MR effects in the plasma-based analyses, resulting in 587 protein-IDP-disease combinations for MVMR testing.

Across 12 neurological and psychiatric disorder GWAS, we identified 22 unique plasma protein-disease pairs with significant total effects (Benjamini-Hochberg FDR < 0.05); however, none showed evidence of significant mediation through IDPs. These associations involved eleven proteins (coding genes *APOE*, *ARL3*, *MICB*, *NSF*, *RHOC*, *RSPO3*, *ENPP2*, *BTN2A1*, *EIF2AK3*, *MRVI1*, and *OPLAH*) and spanned multiple conditions, including Alzheimer’s disease (AD), amyotrophic lateral sclerosis (ALS), bipolar disorder (BD), glioma, major depressive disorder (MDD), multiple sclerosis (MS), Parkinson’s disease (PD), and schizophrenia (**Figure 3; Supplementary Tables 10-11**).

**Figure 3.**
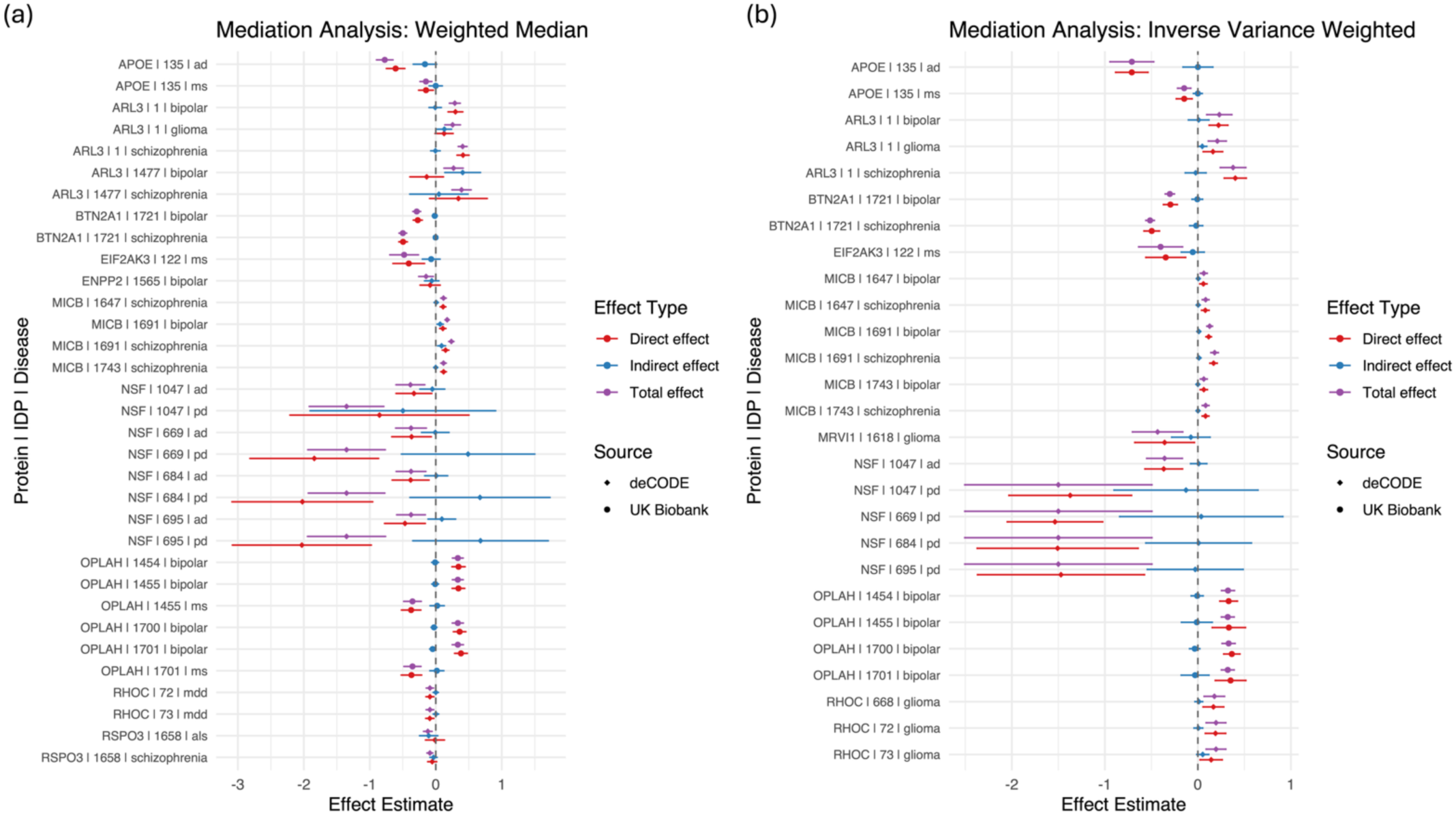
Causal mediation analysis of plasma protein effects on neurological and psychiatric diseases through IDPs using multivariable Mendelian randomization. Plots display the estimated total, direct, and IDP-mediated effects of plasma proteins on disease risk. (a) Weighted median (WM) based MVMR results, showing robust estimates of the total protein effects along with direct and IDP-mediated effects. (b) Inverse-variance weighted (IVW) MVMR results for the same protein-disease pairs. In both panels, the mediated effect represents the proportion of the protein-disease association attributable to variation in the IDP, whereas the direct effect captures the remaining association after accounting for mediation.

### Implicating Brain Cell Types in Plasma Protein Regulation using Single-Cell eQTLs

To further elucidate the cellular context underlying the disease-associated plasma proteins identified through MVMR, we performed MR analyses using single-cell eQTL instruments derived from human brain tissue. Specifically, we assessed whether cell-type-specific gene expression influences plasma protein levels, using plasma protein pQTL summary statistics from the UKB-PPP and deCODE datasets as outcomes. This analysis incorporated 16 single-cell eQTL datasets spanning eight major brain cell types, including astrocytes, endothelial cells, excitatory neurons, inhibitory neurons, microglia, oligodendrocytes, oligodendrocyte precursor cells, and pericytes. For each cell type, eQTLs were sourced from either (i) disease-free control samples or (ii) a combined cohort including both controls and individuals with Alzheimer’s disease, multiple sclerosis, or Parkinson’s disease, as reported in Haglund *et al*.

2025.^19^

We examined all eleven plasma proteins that showed significant total effects in the MVMR analysis. With the exception of *MRVI1*, which lacked SNP instruments, each protein was assessed across both control-only and full cohorts (**Supplementary Table 12**), yielding 151 protein-cell-type-cohort combinations. Of these, 81 associations reached Bonferroni-corrected significance across brain cell types (**Figure 4**). Notably, results obtained from the full case-control cohort showed enrichment patterns that differed from those observed in the disease-free control cohort, suggesting that disease state may moderate cell-type-specific regulatory influences on plasma protein expression.

**Figure 4.**
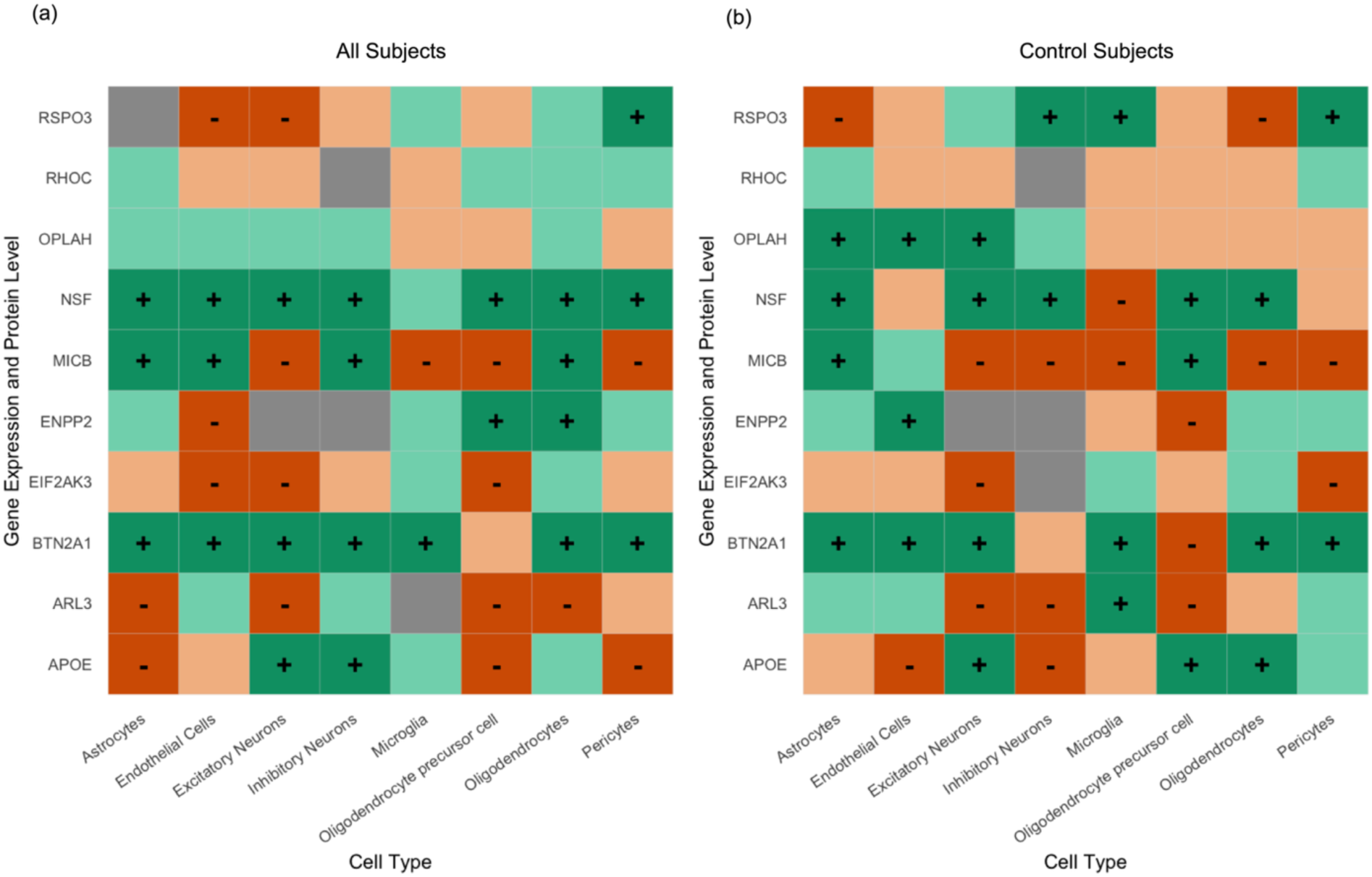
Summary of brain single-cell cis-eQTL effects on plasma protein levels. Heatmaps display the Mendelian randomization-derived associations between cell-type-specific gene expression and corresponding plasma protein levels for the full cohort (A) and control-only samples (B). Colors and symbols indicate the direction and statistical strength of the associations: dark green with a plus (+) symbol denotes a significant positive association, while light green indicates a non-significant positive trend. Dark orange with a minus (–) symbol denotes a significant negative association, and light orange represents a non-significant negative trend. Gray shading indicates protein-cell type combinations without a valid single-SNP instrument.

### Characterizing Imaging-Related Biological Pathways

Given the absence of evidence that variation in brain structure mediates disease risk, we interpret the disease-associated proteins as upstream factors that independently manifest in both IDPs and disease outcomes. We therefore sought to characterize biological processes that might jointly explain variation in plasma protein levels and their corresponding IDPs. To this end, we first computed gene-level association statistics for each IDP GWAS using MAGMA and subsequently performed pathway enrichment analyses on the resulting gene sets using Enrichr.^20,21^ This analysis revealed multiple convergent biological processes implicated in IDP variation, many of which were concordant with known functions of the associated plasma proteins (**Supplementary Table 13**).

Several notable pathway-disease relationships emerged. Ion transport, ferroptosis, and insulin resistance pathways involving APOE were enriched in IDPs associated with Alzheimer’s disease (AD) and multiple sclerosis (MS). Wnt signaling, vesicle trafficking, and neurogenesis linked to NSF were enriched in IDPs relevant to Parkinson’s disease (PD) and AD. Immune signaling, antigen presentation, and T cell and natural killer cell cytotoxicity, mediated by *MICB* and *BTN2A1*, mapped to IDPs associated with bipolar disorder (BD) and schizophrenia. Hedgehog signaling and epigenetic chromatin regulation related to *ARL3* appeared in IDPs implicated in BD, schizophrenia, and glioma. Pathways related to human papillomavirus infection and extracellular matrix involving *MRVI1* were enriched in IDPs linked to glioma. Actin dynamics and morphogen signaling associated with *RHOC* were observed in IDPs relevant to glioma and major depressive disorder (MDD). Cytoskeletal and intermediate filament pathways implicated by *OPLAH* (BD and MS) and RSPO3 (ALS and schizophrenia), as well as antigen presentation and stress pathways involving *EIF2AK3* in MS provided further mechanistic support. Collectively, these findings highlight biologically coherent pathways through which plasma protein variation may contribute to the observed diversity in brain structural phenotypes, complementing their independent effects on neurological and psychiatric disease risk.

### Prospective Drug Interactions

Druggable proteins identified through the MVMR analysis (**Supplementary Table 14**) showed strong drug-target interactions for eight out of the eleven prioritized proteins: *APOE* (n = 58), *ARL3* (n = 7), *RHOC* (n = 6), *ENPP2* (n = 6), *MICB* (n = 3), *RSPO3* (n = 1), *EIF2AK3* (n = 1), and *OPLAH* (n = 1). These protein-drug relationships provide mechanistic insight into disease-associated pathways and highlight therapeutic opportunities for modulating plasma protein-related biology. Interactions between *ARL3* and *RHOC* and several hormonal agents suggest potential links to neuroendocrine signaling processes implicated in BD, MDD, and glioma. Interactions involving *MICB* and *RSPO3* with immune-modulating or antiviral therapies point to immunological and inflammatory mechanisms contributing to schizophrenia and amyotrophic lateral sclerosis (ALS). Targeting of *ENPP2* and *EIF2AK3* by metabolic or stress-response modulators implicates involvement of cellular metabolism and stress signaling in BD and MS. Together, these results, summarized for all eleven prioritized proteins in **Table 2** and **Supplementary Table 15**, underscore the therapeutic relevance of plasma protein dysregulation and outline multiple avenues for future drug development.

**Table 2:**
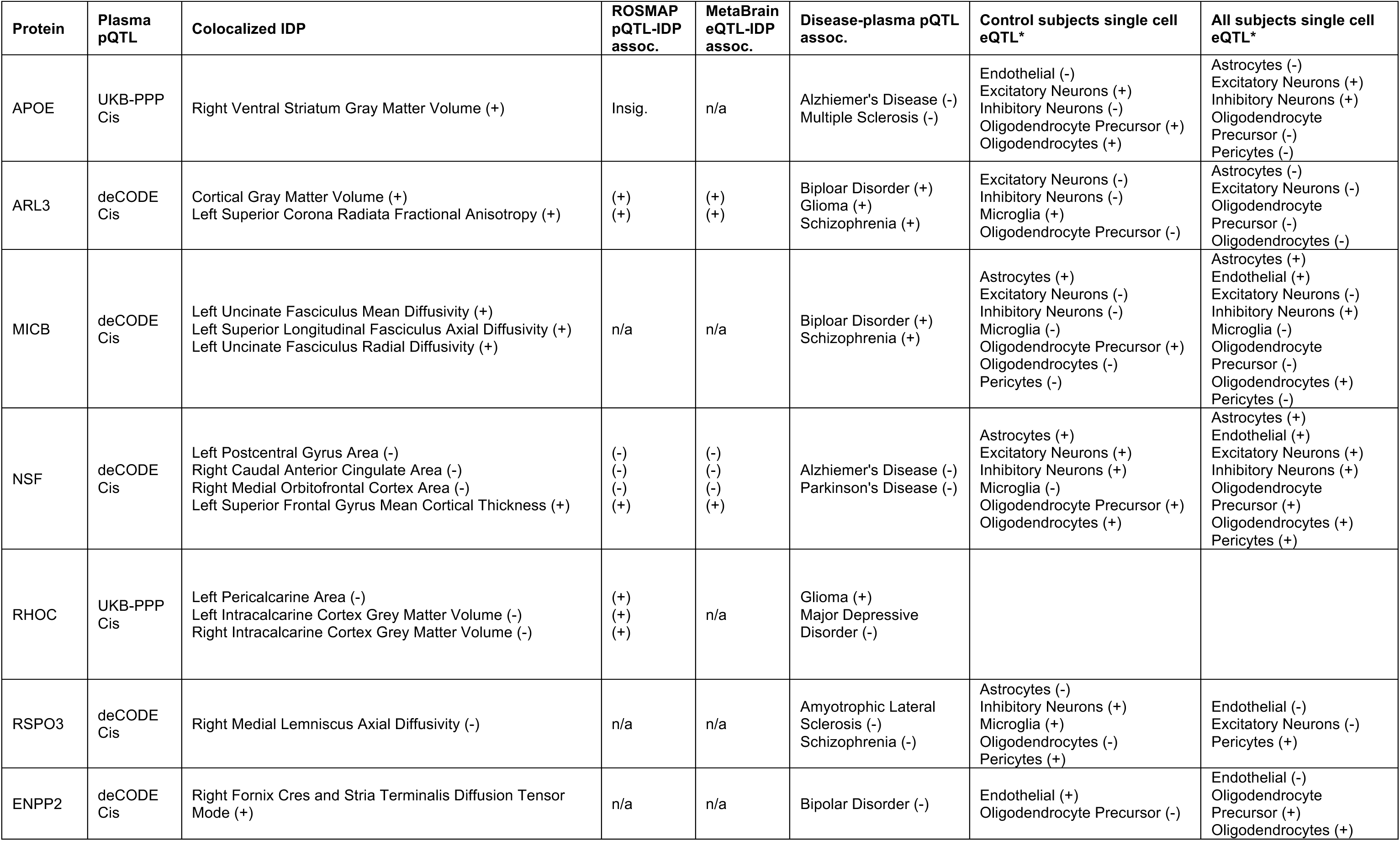
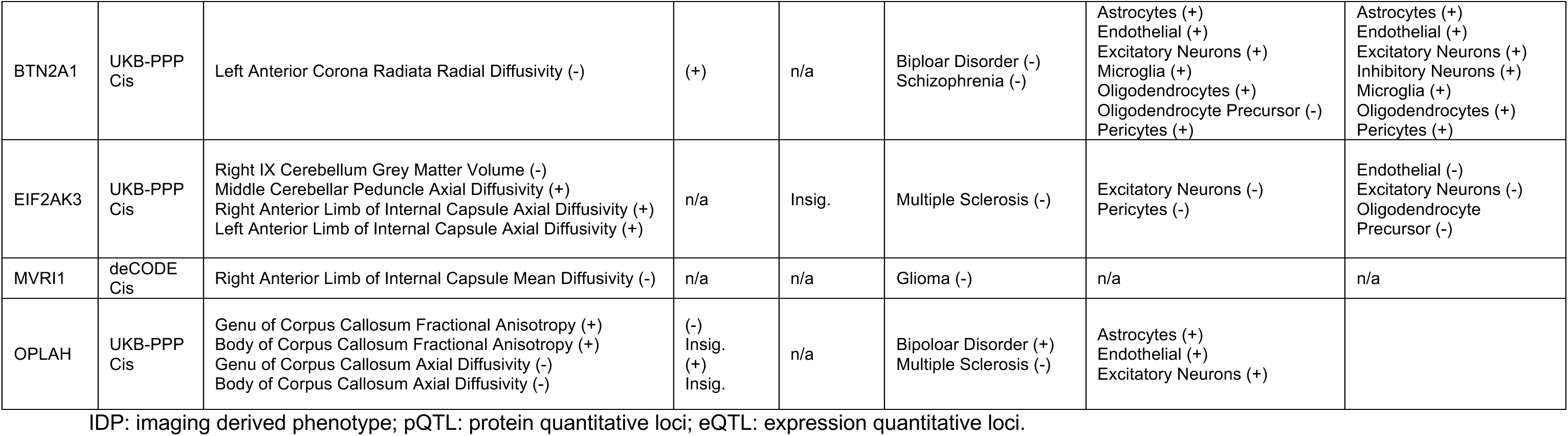
Summary of evidence for proteins and genes identified through Multivariable Mendelian Randomization.

## Discussion

In this study, we integrated the two largest plasma pQTL resources to characterize how genetically regulated plasma protein levels relate to population-level variation in brain IDPs. Through MR and colocalization analyses, we identified 120 cis-pQTL and 20 trans-pQTL associations with IDPs, many of which were further validated using brain-tissue-derived proteomic and transcriptomic datasets. These results underscore the biological relevance of plasma-derived signals and highlight shared mechanisms between circulating protein regulation and brain structure.

Several cis-pQTL findings mapped to proteins influencing multiple brain regions. *BCAN*, which encodes brevican, an extracellular matrix component enriched in perineuronal nets, was associated with structural variation in several major white-matter tracts, including the corona radiata, corpus callosum, and longitudinal fasciculus. These associations align with the established role of brevican in synaptic plasticity^22^, neuronal development, and synaptic remodeling.^23^ Similarly, *FKBPL*, a modulator of glucocorticoid signaling with known functions in angiogenesis and inflammation, was associated with white-matter organization in the posterior thalamic radiation, sagittal stratum, and internal capsule.^24^ Neither protein showed direct effects on disease risk, suggesting that some protein-IDP associations reflect normative structural variability rather than pathogenesis.

Among plasma proteins associated with brain structure, eleven also demonstrated significant effects on neurological or psychiatric disorders. Interestingly, these effects were direct rather than mediated through IDPs, indicating that protein influences on brain structure and disease may represent parallel causal pathways. Enrichment analyses of IDP-associated genes revealed biological processes consistent with known functions of implicated proteins. Single-cell analyses further mapped these effects to specific brain cell populations, providing additional mechanistic insight.

For instance, plasma APOE abundance exhibited a protective association with AD, consistent with prior reports.^25^ Cell-type-specific patterns were most prominent in inhibitory neurons and oligodendrocyte precursor cells, aligning with established *APOE*-related biology.^26^ *APOE* was strongly associated with ventral striatal gray-matter volume, and IDP-enriched pathways implicated metal ion transport, a process previously linked to AD pathophysiology.^27^ Although these converging molecular and imaging signals are suggestive, it remains uncertain whether structural changes mediate disease risk or reflect parallel consequences of APOE dysregulation.

*NSF*, a central membrane trafficking protein, demonstrated widespread associations with cortex thickness in prefrontal and cingulate regions, areas known to be vulnerable in AD.^28^ Our analysis also replicated NSF associations with PD, consistent with its involvement in dopaminergic-related pathogenesis.^29^ Single-cell analyses implicated both excitatory and inhibitory neurons, and IDP gene enrichment highlighted pathways related to synaptic vesicle dynamics and WNT signaling, two processes central to neuronal homeostasis.^30^ The convergence of AD- and PD-related genetic signals in NSF-associated IDPs suggests distinct but partially overlapping mechanisms affecting both structural variation and disease risk.

The effects of *MICB* and *BTN2A1* localized to frontotemporal white-matter tracts, including the anterior corona radiata and uncinate and superior longitudinal fasciculi, regions implicated in mood and psychotic disorders.^31,32^ Functional annotations revealed enrichment for MHC, interferon, and nuclear remodeling pathways, consistent with the immunological functions of *MICB* and *BTN2A1* in natural killer and T cell activities.^33,34^ These findings reinforce neuroimmune dysfunction as a core mechanism linking white-matter microstructure to the risk of schizophrenia and BD.^35^

*ARL3*, a regulatory GTPase involved in ciliary signaling and microtubule trafficking, was associated with BD and schizophrenia and has been implicated across diverse ancestral populations.^36^ Its associations with peripheral cortical gray matter and corona radiata, combined with enrichment for hedgehog signaling, support a neurodevelopmental mechanism. *OPLAH*, a critical enzyme in glutathione metabolism, was linked to central callosal white matter and showed enrichment for pathways related to systemic lupus erythematosus, an autoimmune condition associated with increased BD risk and overlapping neuropsychiatric manifestations.^37,38^

In demyelinating disorders such as MS, our findings highlight both *EIF2AK3* and *OPLAH* as plasma proteins contributing to disease risk. *EIF2AK3*, a core mediator of endoplasmic reticulum stress response, has been repeatedly implicated in MS.^39,40^ Thalamic IDPs associated with *EIF2AK3* showed enrichment for stress response and endoplasmic reticulum related pathways, consistent with *EIF2AK3*-medicated neurodegeneration. *OPLAH*’s association with callosal white matter aligns with established MS pathology involving corpus callosum demyelination.^41^ Enrichment of neutrophil extracellular trap formation and oxidative stress pathways further emphasizes a shared cascade.^42^

In glioma, significant associations were identified for plasma *RHOC*, *ARL3*, and *MRVI1*. *RHOC* regulates cytoskeletal remodeling and cell motility, while *ARL3* controls ciliary trafficking and microtubule-dependent transport, both processes relevant to tumor progression.^43,44^ Their associations with intracalcarine and peripheral cortical gray matter measures suggest effects on developmental pathways. Notably, IDP-enriched pathways showed limited overlap with glioma-specific biology, except for hedgehog signaling in peripheral cortical gray matter, implying that these structural associations may reflect developmental rather than tumor-specific mechanisms.

While this study provides a large-scale, integrative evaluation of plasma protein influences on brain structure and disease, several limitations merit discussion. First, the two-sample MR design assumes no sample overlap between exposure and outcome datasets. Although the deCODE pQTL dataset fully satisfies this assumption, UKB-PPP shares ∼9% sample overlap with UKB imaging GWAS. However, recent study suggested that the bias caused by sample overlap in two-sample MR analysis is minimal.^45^ Additionally, given that we utilized strong genetic instruments from large-scale pQTL datasets and successful replication across independent datasets, any potential biases due to sample overlap is expected to be small.

Second, despite leveraging the largest proteomic and imaging datasets available, statistical power remains constrained, particularly for mediation analyses, which are known to be underpowered.^46^ Third, replication using brain tissue derived pQTL and eQTL datasets remains limited by modest samples sizes (n = 706 for brain pQTLs; n = 6,526 for brain eQTLs; n = 183 controls and n = 391 total subjects for brain single-cell eQTLs). Some non-replicating associations may thus reflect insufficient power rather than absence of biological effect. Fourth, we used the UKB-PPP LD reference panel for deCODE analyses due to lack of an Icelandic reference panel, which may introduce LD mismatch that reduces power and increases bias.

Despite these limitations, our integrative multi-omics framework connects plasma protein variation to brain structural phenotypes and disease risk, revealing both convergent and distinct biological pathways. By cross-validating plasma-derived associations with brain proteomic, transcriptomic, and single-cell data, we identify proteins with potential mechanistic roles in brain structure, function, and disease susceptibility. Our findings indicate that circulating protein effects on disease risk may be independent of their effects on brain structure and these effects may manifest in specific anatomical regions and cell types. By pinpointing these anatomical and cellular contexts, our work refines mechanistic models of neurobiological variation and highlights plasma proteins as accessible markers and potential therapeutic targets. Future mechanistic and translational research in disease-specific cohorts will be critical for validating causal pathways and advancing precision medicine applications.

## Methods

### Plasma Proteomic Quantitative Trait Loci

We utilized plasma pQTL data from two large-scale studies: Sun *et al.* 2023 (UKB-PPP) and Ferkingstad *et al.* 2021 (deCODE).^5,6^ The UKB-PPP dataset provided pQTL summary statistics for 2,922 plasma proteins measured in 34,557 individuals of European ancestry in the discovery cohort using the antibody-based Olink Explore 3072 proximity extension assay.^6^ Summary statistics were obtained from the Synapse repository (https://www.synapse.org/Synapse:syn51364943/wiki/622119). The deCODE dataset included pQTL data for 4,907 aptamers corresponding to 4,719 proteins measured in 35,559 Icelandic individuals of European ancestry, generated using the SomaScan multiplex aptamer assay version 4.^5^ Summary statistics were downloaded from the deCODE repository (https://www.decode.com/summarydata/). Additional methodological details for both pQTL studies are available in the original publications.^5,6^

We identified genetic instruments for Mendelian randomization (MR) analysis following the procedures described in Bhattacharyya *et al.* 2025.^47^ pQTL GWAS summary statistics underwent quality control (QC), including removal of indels, variants with minor allele frequency (MAF) <0.001, palindromic variants with MAF >0.42, and specifically for trans-pQTLs, variants within the extended MHC region (Chr6:25-34Mb). Filtered datasets were converted to GWAS VCF format using the gwas2vcf tool,^48^ and genome-wide significant variants were clumped to identify independent genetic instruments. Variants located within 1 Mb of the protein-coding gene transcription start site were classified as cis-pQTLs, whereas variants outside this window were classified as trans-pQTLs. LD clumping was performed separately for cis- and trans-pQTLs using the IEUGWASR tool^49^ with the UKB-PPP samples of European ancestry as the LD reference panel.

### Genome-Wide Association Studies of Brain Imaging Derived Phenotypes

We obtained GWAS summary statistics for brain IDPs from 33,224 UKB participants of European ancestry, as reported by Smith *et al.* 2021.^14^ The full dataset included 3,935 IDPs spanning across six MRI modalities: T1-weighted structural MRI, T2-weighted FLAIR, susceptibility-weighted MRI, diffusion MRI, task functional MRI, and resting-state functional MRI. For the present study, we focused on 556 distinct and representative IDPs derived from T1-weighted and diffusion MRI modalities, selecting phenotypes that capture key neuroanatomical and microstructural features (**Supplemental Table 2**). Each IDP GWAS included 17,103,079 genetic variants. We performed a genome build liftover from GRCh37 to GRCh38 reference genome, resulting in 17,100,178 variants retained for downstream analyses. Further details regarding imaging processing and GWAS methods are available in Smith *et al.* 2021.^14^

### Brain Tissue-Derived Proteomics Quantitative Trait Loci

To validate the plasma pQTL-IDP associations, we utilized brain tissue derived pQTL summary statistics generated from the ROSMAP cohort by Wingo *et al.* 2023.^18^ The study analyzed human brain proteomes from the dorsolateral prefrontal cortex of 706 postmortem samples from individuals of European ancestry. We applied the same quality control and processing steps as for the plasma pQTL instruments, retaining only variants with a genotyping calling rate above 95%. For each protein, we selected the single most significant pQTL as the genetic instrument. The resulting instrument set included 5,259 distinct proteins for downstream analysis. We then restricted this set to a replication subset, retaining only proteins that were statistically significant in either the primary plasma cis-pQTL or trans-pQTL MR analyses. Further details on the ROSMAP brain pQTL generation and processing are available in the original publication.^18^

### Brain Tissue-Derived Expression Quantitative Trait Loci

To further validate associations identified in the primary plasma pQTL analysis, we leveraged brain tissue-derived eQTL data from the MetaBrain consortium^2^, which aggregated bulk RNA-seq and genotype data across multiple studies, providing a comprehensive brain eQTL resource. The summary statistics used in this study were generated from 8,613 samples across six brain regions, with the cortex contributing the largest sample size (n = 6,526). 16,169 cis-eQTLs were identified in the European ancestry cortex samples. We applied the same quality control and processing pipeline used in the ROSMAP pQTL analysis, selecting the single most significant eQTL as the genetic instrument for each gene. This resulted in 11,256 instruments, which we further restricted to genes corresponding to proteins identified as significant in the primary plasma cis- or trans-pQTL MR analysis. Additional details regarding the MetaBrain study are available in the original publication.^2^

### Brain Single-Cell Expression Quantitative Trait Loci

For the single-cell analyses, we used brain single-cell cis-eQTL data spanning eight major brain cell types – excitatory neurons, inhibitory neurons, astrocytes, microglia, oligodendrocytes, oligodendrocyte precursor cells (OPCs), endothelial cells, and pericytes – reported in Haglund *et al.* 2025.^19^ Drawing on four previously published studies, Haglund *et al.* generated a comprehensive single-cell RNA sequencing and genotyping dataset from postmortem brain tissue samples of individuals of European ancestry. The brain single-cell cis-eQTL data were derived from 391 samples, including 183 neurologically healthy controls and 208 individuals diagnosed with brain disorders Parkinson’s disease (PD), multiple sclerosis (MS), or Alzheimer’s disease (AD). In total, the dataset encompassed 2,348,438 single nuclei, yielding approximately 1.82 million brain single-cell cis-eQTLs (FDR < 5%) mapped to 13,939 unique genes. For our analyses, we used 16 single-cell eQTL datasets corresponding to the eight cell types, each generated separately from (i) control-only samples and (ii) the combined cohort including disease and control samples. Additional information regarding data collection and processing are available in the original publication.^19^

### Mendelian Randomization

MR analyses were conducted using the TwoSampleMR package (version 0.6.6) in R (version 4.2.2). SNPs significantly associated with molecular phenotypes, including plasma protein levels, brain gene expression, brain protein abundance, and single-cell gene expression, were used as instrumental variables to estimate the causal effects of each protein on IDPs or disease outcomes. For single-instrument analyses, causal estimates were derived using the Wald ratio. When at least two instruments were available, we performed inverse-variance weighted (IVW) MR, and when three or more instruments were available, we additionally conducted weighted median (WM) MR using default settings. Effect estimates (beta) were made robust to pleiotropic SNPs through penalization (constant = 20), robust estimation using the Huber loss function, and a scale factor of 1. Confidence intervals and standard errors were adjusted for overdispersion, assuming no covariance between SNP-exposure and SNP-outcome associations. Statistical inference was based on two-sided z-score tests with heterogeneity assessed using a 5% significance threshold.

For the primary plasma pQTL analyses, we used a hierarchical approach to minimize false positives due to pleiotropy, prioritizing MR methods in the following order: WM (most robust to pleiotropic bias), followed by IVW, and finally single-SNP Wald ratio estimates.^9^ MR analyses were conducted separately for each pQTL dataset, and statistical significance was determined using Bonferroni-corrected thresholds reflecting the number of proteins and IDPs tested (UKB-PPP cis-pQTL: *P* < 4.48×10^−8^; UKB-PPP trans-pQTL: *P* < 3.54×10^−8^; deCODE cis-pQTL: *P* < 5.41×10^−8^, deCODE trans-pQTL: *P* < 1.91×10^−8^). Because trans-pQTL instruments arise from dispersed genomic regions and are thus more susceptible to pleiotropic bias, only WM MR results were considered for trans-pQTL analyses. In the ROSMAP and MetaBrain validation analyses, each protein or gene was instrumented by a single SNP, and thus Wald ratio estimates were used. Similarly, for all single-cell analyses, we limited instruments to the most significant SNP per gene.

### Colocalization

Following the primary plasma MR analysis, we performed colocalization analysis using the coloc R package (version 5.2.2) to evaluate whether significant cis-pQTL-IDP associations arose from a shared causal variant. This secondary analysis helped distinguish true shared genetic signals from associations driven by LD. For each locus, coloc evaluates five mutually exclusive hypotheses: H0: no association with either trait; H1: association with trait 1 only; H2: association with trait 2 only; H3: two distinct causal variants; H4: a single shared causal variant. Colocalization was performed using SNPs within 1.5 Mb of the transcription start site of each protein-coding gene. Prior probabilities for association with trait 1 (p1) and trait 2 (p2) were set to 0.0001, and the prior for a shared association (p12) to 0.00001. Posterior probabilities for the five hypotheses were computed from the observed summary statistics.

The standard coloc method assumes a single causal variant per trait, which may not hold in regions harboring multiple independent signals. To address this limitation, we complemented the standard coloc approach (coloc.abf) with a fine-mapping based colocalization method, SuSiE-coloc, which accounts for multiple causal variants within a region.^17^ Although SuSiE-coloc relaxes the single causal variant assumption, it relies on successful fine-mapping for both traits. For this reason, we used coloc.abf and SuSiE-coloc as complementary methods. Associations were considered colocalized if the posterior probability for H4 exceeded 0.7 in either colocalization method.

### Multivariable Mendelian Randomization Mediation Analysis

We performed mediation analyses within the MR framework to quantify the extent to which plasma protein levels influence neurological and psychiatric disorders through their effects on brain IDPs. Using both cis- and trans-pQTL instruments, along with IDP instruments generated by the same processing pipeline described above, we implemented multivariable Mendelian randomization (MVMR).^46^ To reduce pleiotropic bias and mitigate violations of instrumental variable assumptions, we applied the difference MVMR estimators (Diff-IVW and Diff-Median) with robust outcome formulations. Under a linear mediation model, these methods enabled estimation of total, direct, and indirect (IDP-mediated) effects.

We restricted MVMR analyses to protein-IDP pairs that showed either significant evidence of colocalization or significant trans-pQTL WM MR effects. Each MVMR model was structured with plasma protein concentration as the primary exposure, the corresponding IDP as the mediator, and neurological or psychiatric diseases as outcomes. Disease outcomes were drawn from the largest publicly available European-ancestry GWAS meta-analyses, including AD (EADB, GRACE, FinnGen; 90,338 cases / 1,036,225 controls), PD (IPDGC; 37,688 cases and 18,618 proxy cases / 1,417,791 controls), ischemic stroke (IS), cardioembolic ischemic stroke (CIS), small vessel ischemic stroke (SVIS), and large vessel ischemic stroke (LVIS) (GIGASTROKE; 110,182 cases / 1,503,898 controls), glioma (GliomaScan; 12,496 cases / 18,190 controls), ALS (Project MinE; 29,612 cases / 122,656 controls), MS (IMSGC; 47,429 cases / 68,374 controls), schizophrenia (PGC Phase 3; 76,755 cases / 243,649 controls), BD (PGC Phase 3; 41,917 cases / 371,549 controls), and MDD (PGC, MVP, FinnGen; 371,194 cases / 978,693 controls).^47,50–55^

Note that, given concern for exposure-outcome overlap with the UK Biobank, the PD and stroke GWAS were exclusively run with deCODE proteomics data and not the UKB-PPP. Moreover, the schizophrenia, BD and MDD data were derived from Bhattacharyya *et al.* 2025,^47^ which did not include 23andMe data and removed UK Biobank contributions. Because some outcome summary statistics lacked reported effect-allele frequencies (specifically MS and glioma), we imputed allele frequencies by averaging the corresponding frequencies from the plasma protein instruments and the IDP GWAS, leveraging the fact that all databases were derived from European ancestry samples. Results for Diff-IVW and Diff-Median were interpreted separately due to their differing methodological assumptions. Statistical significance was defined as a Benjamini-Hochberg FDR < 0.05 for the total effect within each disease.

### Single-Cell eQTL-based Mendelian Randomization Analysis

We conducted single-cell eQTL-based MR analyses focusing on plasma proteins that were statistically significant in the preceding MVMR analyses. For each protein, we selected the most significant cis-eQTL instrument for the corresponding gene from each brain single-cell eQTL dataset reported in Haglund *et al.*, 2025, reflecting the limited sample size of the single-cell datasets. Wald ratio MR analyses were then conducted for each gene-protein pair, harmonizing alleles and effect directions between the single-cell eQTL instruments (exposure) and the plasma pQTL summary statistics (outcome). Analyses were conducted separately for each combination of plasma protein GWAS dataset (UKB-PPP or deCODE) and each single-cell sample set (control-only or complete cohort). Significant MR associations, representing links between plasma protein abundance and gene expression in specific brain cell types, were identified across both control and complete cohorts, using Bonferroni-corrected thresholds appropriate to each dataset (UKB-PPP Control: *P* < 0.05/31 = 1.61×10^−3^; UKB-PPP Complete: *P* < 0.05/32 = 1.56×10^−3^; deCODE Control: *P* < 0.05/45 = 1.11×10^−3^; deCODE Complete: *P* < 0.05/43 = 1.16×10^−3^). Of the 160 protein-cell type test combinations, nine were excluded due to missing outcome summary statistics for the corresponding single-SNP instrument.

### Biological Pathway Enrichment

We performed pathway enrichment analyses on genes associated with IDPs that formed significant plasma protein-IDP pairs in the MVMR analysis. Gene-level association testing for each IDP GWAS was conducted using MAGMA (v1.10).^20^ For each IDP, SNPs were mapped to genes using the --annotate option with the GRCh37 reference genome and an annotation window extending 35 kb upstream and 10 kb downstream of the transcription start and end sites. Gene-level p-values were computed using MAGMA’s SNP-wise mean model, with LD estimated from the 1,000 Genomes European samples. Enrichment of IDP-associated gene sets was performed using the Enrichr package (version 3.4)^21^ in R (version 4.2.2) across four biologically relevant pathway databases: GO_Biological_Process_2025, GO_Cellular_Component_2025, KEGG_2021_Human, and Reactome_Pathways_2024. These analyses were used to evaluate convergence or divergence of biological processes implicated by the IDP GWAS results. Multiple testing was controlled using the Benjamini-Hochberg procedure, with significance defined as FDR <0.05.

### Druggability Analysis

We assessed the druggability of proteins identified as statistically significant in the MVMR analyses using DGIdb4.0 (www.dgidb.org). DGIdb integrates expert-curated annotations and text-mined evidence to catalog drug-gene interactions from resources such as DrugBank, PharmGKB, ChEMBL, and Drug Target Commons. It also classifies genes as potentially druggable based on their inclusion in druggable pathways, molecular function categories, and gene families, leveraging sources such as the Gene Ontology, the Human Protein Atlas, and established druggable genome reference lists. Using DGIdb4.0, we queried each protein for known or predicted drug interactions. The search identified approved compounds, immunotherapies, and experimentally characterized chemical agents with reported activity against these protein targets.

## Data availability

All analyses in this study were based on previously published summary-level data or de-identified data from public resources. UK Biobank Pharma Proteomics Project summary statistics for Olink Explore plasma proteins (Sun *et al.* 2023) were obtained from Synapse (project ID syn51364943; https://www.synapse.org/#!Synapse:syn51364943). The deCODE plasma proteomics summary statistics for SomaScan plasma proteins (Ferkingstad *et al.* 2021) were downloaded from the deCODE Genetics summary data portal (https://www.decode.com/summarydata/). UK Biobank imaging-derived phenotype GWAS summary statistics (Smith *et al.* 2021) were downloaded from GWAS Catalog (study accession numbers GCST90002426 to GCST90006360) and descriptions of the GWAS can be found at https://open.oxcin.ox.ac.uk/ukbiobank/big40/. ROSMAP dorsolateral prefrontal cortex proteomics brain pQTL summary statistics (Wingo *et al.* 2023) were accessed through the AMP-AD Knowledge Portal/Synapse under the ROSMAP program (https://doi.org/10.7303/syn51150434; https://adknowledgeportal.synapse.org/Explore/Programs/DetailsPage?Program=AMP-AD). MetaBrain cortex cis-eQTL summary statistics (de Klein *et al.* 2023) were obtained from https://www.metabrain.nl/. Brain cell-type specific cis-eQTL summary statistics (Haglund *et al.* 2025) were downloaded from https://zenodo.org/records/13343729. Summary statistics for neurological and psychiatric disorder GWAS are available from the respective consortia or public repositories. Reference details are provided below: 1) Alzheimer’s disease: EADB / GRACE / FinnGen meta-analysis (Wightman *et al.* 2021); 2) Parkinson’s disease: IPDGC meta-analysis (Nalls *et al.* 2019); 3) Ischemic stroke and subtypes: GIGASTROKE consortium (Mishra *et al.* 2022); 4) Glioma: GliomaScan (Melin *et al.* 2017); 5) Amyotrophic lateral sclerosis: Project MinE (van Rheenen *et al.* 2021); 6) Multiple sclerosis: IMSGC (International Multiple Sclerosis Genetics Consortium 2019); 7) Schizophrenia, bipolar disorder, and major depressive disorder: PGC Phase 3 and related meta-analyses as implemented in Bhattacharyya *et al.* 2025, excluding UK Biobank and 23andMe contributions.

All summary statistics generated in this study are provided in the supplementary materials of this study or otherwise can be obtained from the corresponding authors upon reasonable request, subject to original data-use agreements.

## Code availability

All statistical analyses were conducted in R (version 4.2.2). The following packages and software were used: TwoSampleMR (version 0.6.6); ieugwasr (version 1.1.0.9000); coloc (version 5.2.2); MAGMA (version 1.10); Enrichr R package (version 3.4).

## Acknowledgement

U.B., J.J., M.L., and T.L. were supported by the National Institute of Mental Health of the National Institutes of Health (NIH) under award no. R01MH117646 (T.L., principal investigator). The funding agency had no direct role in the design and conduct of the study; collection, management, analysis, and interpretation of the data; preparation, review, or approval of the manuscript; and decision to submit the manuscript for publication. The content is solely the responsibility of the authors and does not necessarily represent the official views of the NIH.

## Author contribution

C.A., T.G. and C-Y.C. conceived and designed the study. T.G. and C-Y.C jointly supervised the study. C.A., E.D., U,B., and J.J. performed data curation and quality control. T.L. and M.L supervised the data curation and quality control. C.A. and E.D. performed statistical analyses. C.A. and C-Y.C. wrote the first draft of the manuscript. T.G., M.L., T.L., U.B. and J.J. critically revised the manuscript. All authors reviewed and approved the final version of the manuscript.

