## Supplementary Figures for "Characterizing the impact of plasma protein levels on human brain structure and disorders leveraging integrative multi-omics analysis"

[$]Current address: Merck & Co., Inc., Cambridge, MA, USA

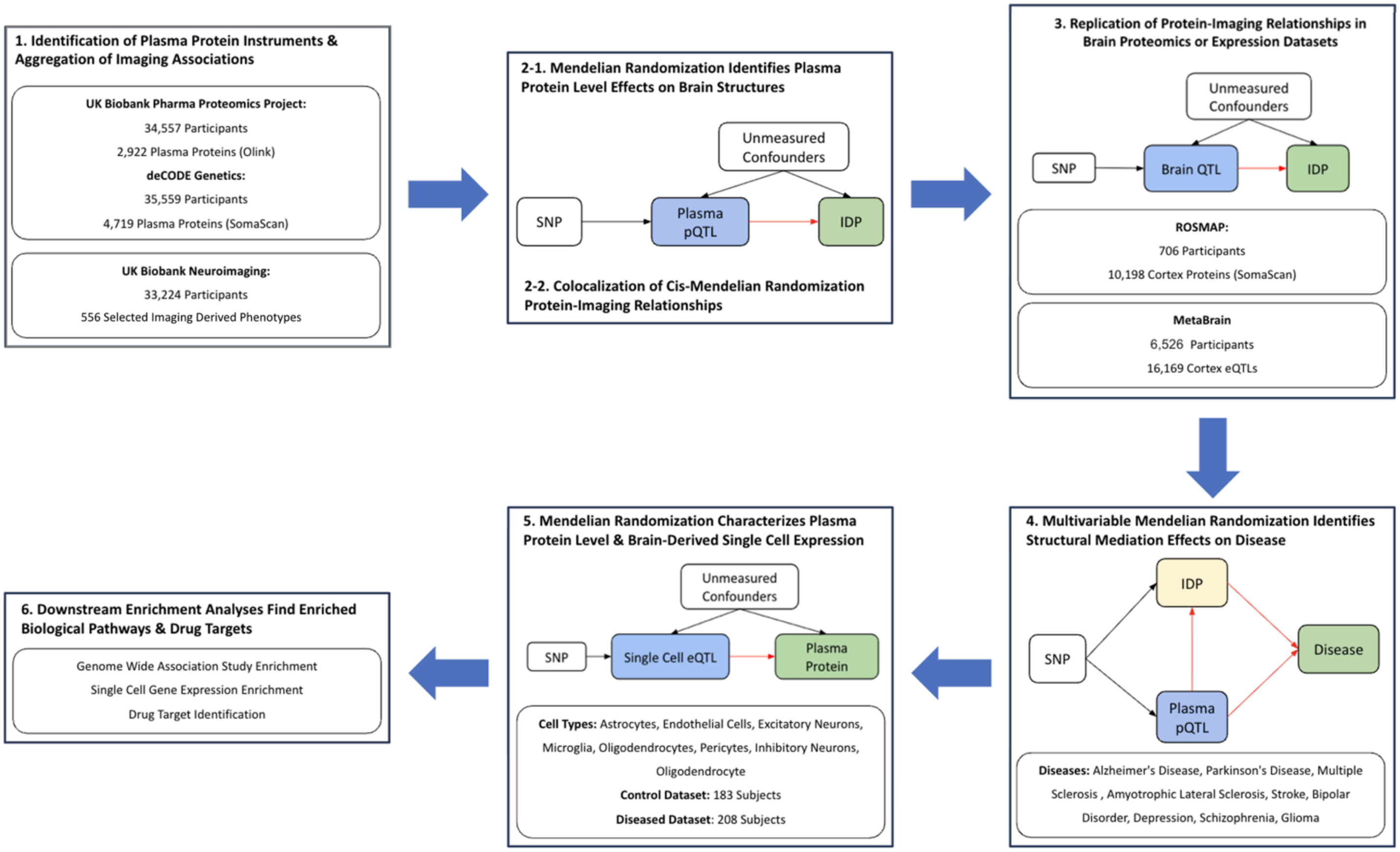


**Supplementary Figure 1. Overview of the analytic framework.**

Overview of the analytic workflow accompanied by the corresponding directed acyclic graphs. Abbreviations: pQTL, protein quantitative trait locus; eQTL, expression quantitative trait locus; SNP: single nucleotide polymorphism; IDP: imaging-derived phenotype; ROSMAP: Religious Orders Study and Memory and Aging Project.

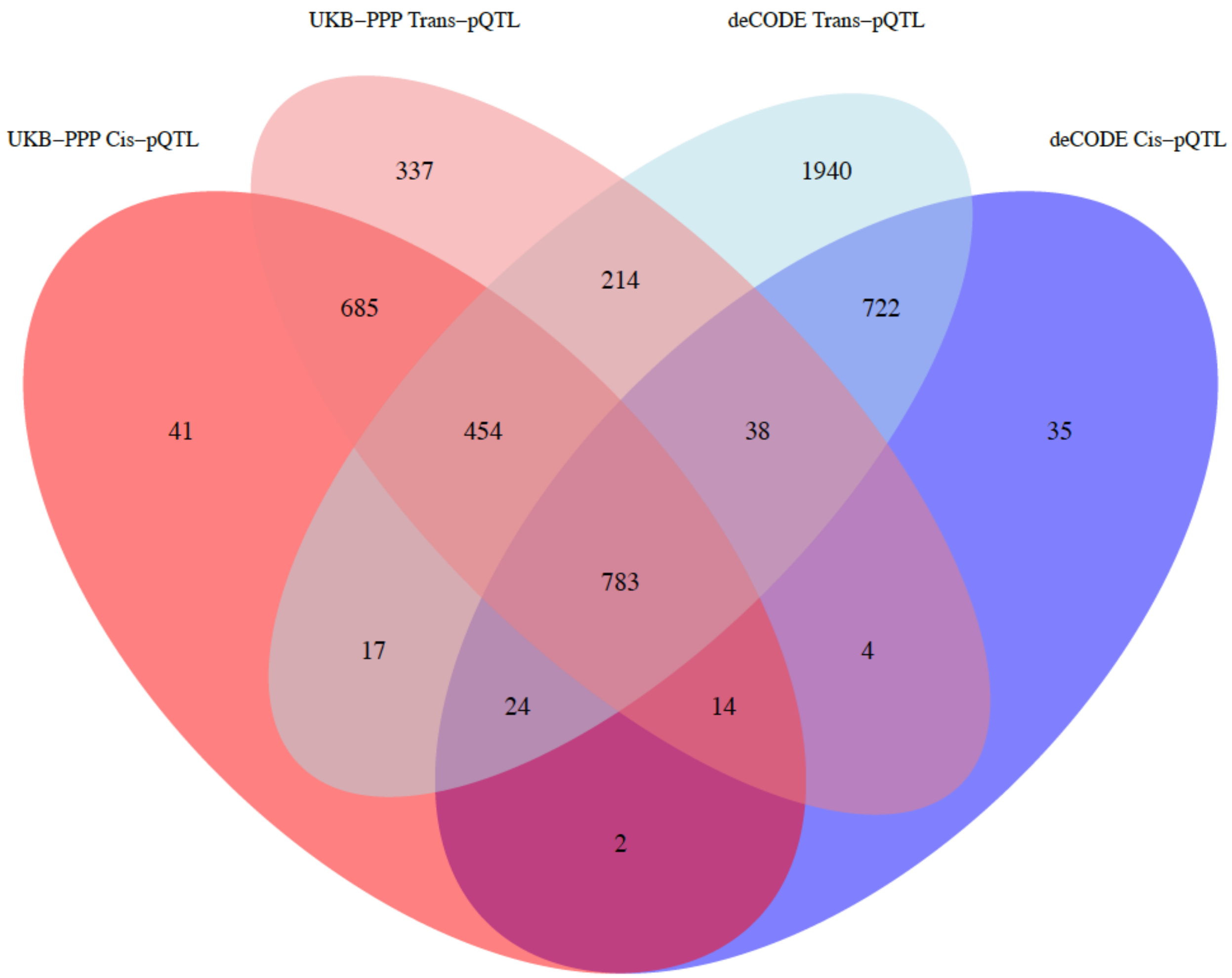


**Supplementary Figure 2. Overlap of proteins with genetic instruments in the UKB-PPP and deCODE datasets after data processing and quality control.**

Each circle in the Venn diagram represents the number of unique plasma proteins with curated genetic instruments (cis- or trans-pQTLs) within the corresponding dataset. Overlapping regions indicate proteins shared across databases. UKB-PPP, UK Biobank Pharma Proteomics Project; pQTL, protein quantitative trait locus.
